# Real-world experience with switching from originator to biosimilar natalizumab

**DOI:** 10.1101/2025.02.05.25320428

**Authors:** Einar August Høgestøl, Åge Winje Brustad, Elisabeth Gulowsen Celius, Martine Meling, Pål Berg-Hansen, Grete Birkeland Kro, Marton König, David J. Warren, Johanna E. Gehin, Nils Bolstad, Gro Owren Nygaard

**Author notes:** Authors contributed equally to this work (Einar August Høgestøl, Åge Winje Brustad).

## Abstract

In January 2024, all persons with multiple sclerosis (pwMS) treated with natalizumab (NTZ) at Oslo University Hospital switched from originator to biosimilar NTZ. We prospectively monitored 39 pwMS during the first year of treatment with biosimilar NTZ and evaluated change in disease activity, side effects, serum NTZ levels, anti-drug antibodies (ADAb), and anti-John Cunningham virus (JCV) antibody levels. Serum NTZ levels and ADAb were measured using in-house assays, while JCV antibody levels were evaluated using Stratify (Biogen) and Immunowell (Sandoz) platforms. One new relapse occurred during the first year and 11 pwMS (28%) reported new side effects after switching to the biosimilar; whereof fatigue, headache, and muscle pain were most frequent. Serum NTZ levels were similar between pwMS on originator (15.1 mg/L, SD 8.9) and biosimilar NTZ (14.9 mg/L, SD 9.0; p = 0.63). We identified ADAb in one pwMS, present both before and after switching. The proportion of pwMS with positive JCV antibody levels increased from 13% in 2023 (Stratify) to 52% in 2024 (Immunowell). Four pwMS discontinued NTZ due to high anti-JCV antibody levels (in the Immunowell assay) in the first 10 months.

## Introduction

Natalizumab (NTZ), a humanised monoclonal antibody against the α4-integrin subunit displayed on circulating leukocytes, was the first approved high-efficacy disease modifying therapy (HE-DMT) for multiple sclerosis (MS) (1). HE-DMTs have provided major improvements to MS long-term outcomes, but they come with significant healthcare costs (2, 3). Biosimilar drugs are nearly identical to the originator biologic drugs and provide more affordable options compared to the expensive originator. While NTZ was very efficient compared to previous MS therapies once it was first made available for persons with MS (pwMS), its use in Norway has been limited because of the high cost, monthly infusions and the risk of severe progressive multifocal leukoencephalopathy (PML) caused by the John Cunningham virus (JCV) (4, 5). Regular testing for anti-JCV antibody levels is part of standard monitoring of NTZ treatment and has reduced the risk of PML (6, 7).

Anti-drug antibodies (ADAb) may adversely affect long-term safety and efficacy of biologic drugs, including NTZ, in pwMS. ADAb can develop in response to repeated exposure, potentially leading to reduced efficacy, hypersensitivity reactions, or an increased risk of adverse effects. Monitoring for ADAb is a critical component of personalised treatment strategies, as it allows clinicians to identify patients who may be at increased risk of adverse outcomes due to ADAb (8).

Tyruko® (Sandoz) is the first approved biosimilar NTZ to the originator drug, Tysabri® (Biogen), used to treat relapsing forms of MS (9). Tyruko was approved in Norway and became available and assigned as the preferred NTZ drug by the authorities on November 1^st^, 2023. All pwMS treated with NTZ at Oslo University Hospital were switched from originator to biosimilar NTZ in January 2024. We prospectively collected information on new disease activity, side effects, serum drug levels, and ADAb before and after switching. Furthermore, we compared the results from the anti-JCV antibody level test results from the Stratify platform, provided by Biogen prior to switching, with the results from the Immunowell platform, provided by Sandoz in the year following the switch. In this paper, we present our first experiences with switching from originator to biosimilar NTZ.

## Materials and Methods

The study was conducted as a prospective observational study from January 2024 to December 2024, in the period when we switched from originator to biosimilar NTZ. Thirty-nine of the 40 pwMS treated with NTZ at Oslo University Hospital provided written, informed consent and participated in the study. All participants were closely monitored during infusions, asked about side effects at each visit, standard blood samples taken every third moth, and followed with yearly clinical consultations and cerebral MRI as part of routine clinical follow-up. All clinical consultations and MRI assessments conducted during the first year after switch were evaluated by an experienced neurologist in December 2024. Neurological function was evaluated using the Expanded Disability Status Scale (EDSS) (10). All reported side effects by pwMS were considered and evaluated by two neurologists. Possible side effects that were not present before the switch but occurred within the first six months afterward were reported. As part of routine follow-up, all pwMS had samples tested for ADAb against NTZ one year after treatment initiation at the Neuroimmunological Laboratory at Haukeland University Hospital (11). If positive for ADAb, treatment was switched to another DMT. We routinely test for anti-JCV antibody levels twice yearly for all pwMS treated with NTZ. The study was approved by the regional ethics committee of South-East Norway (REK 2011-1846A).

### Drug levels

NTZ is usually given at 4-6 week intervals and serum samples for measurement of NTZ levels and ADAb were collected immediately before the next infusion. The first sample was collected prior to the first infusion of biosimilar NTZ in January 2024 and the second sample was collected prior to the second infusion of biosimilar. Furthermore, we collected additional serum samples in pwMS who reported side effects, for evaluation of intervals between infusions, and when the previous results indicated the need for additional samples. The samples were centrifuged and stored at -20 °C before analysis. Serum NTZ (s-NTZ) levels were measured using a validated 3-step time-resolved fluorescence assay automated on the AutoDELFIA (Revvity Inc., Waltham, MA) platform. The assay was performed in streptavidin-coated 96-well plates using biotinylated recombinant human α4β1-integrin as the capture reagent. Active drug binding to the α4β1-integrin solid phase was detected using europium-labelled protein A as the tracer reagent. We report the last s-NTZ levels of originator before the switch and the first s-NTZ levels after switch to the biosimilar (Figure 1), as well as the longitudinal trend of individual s-NTZ levels (Figure 2).

**Figure 1:**
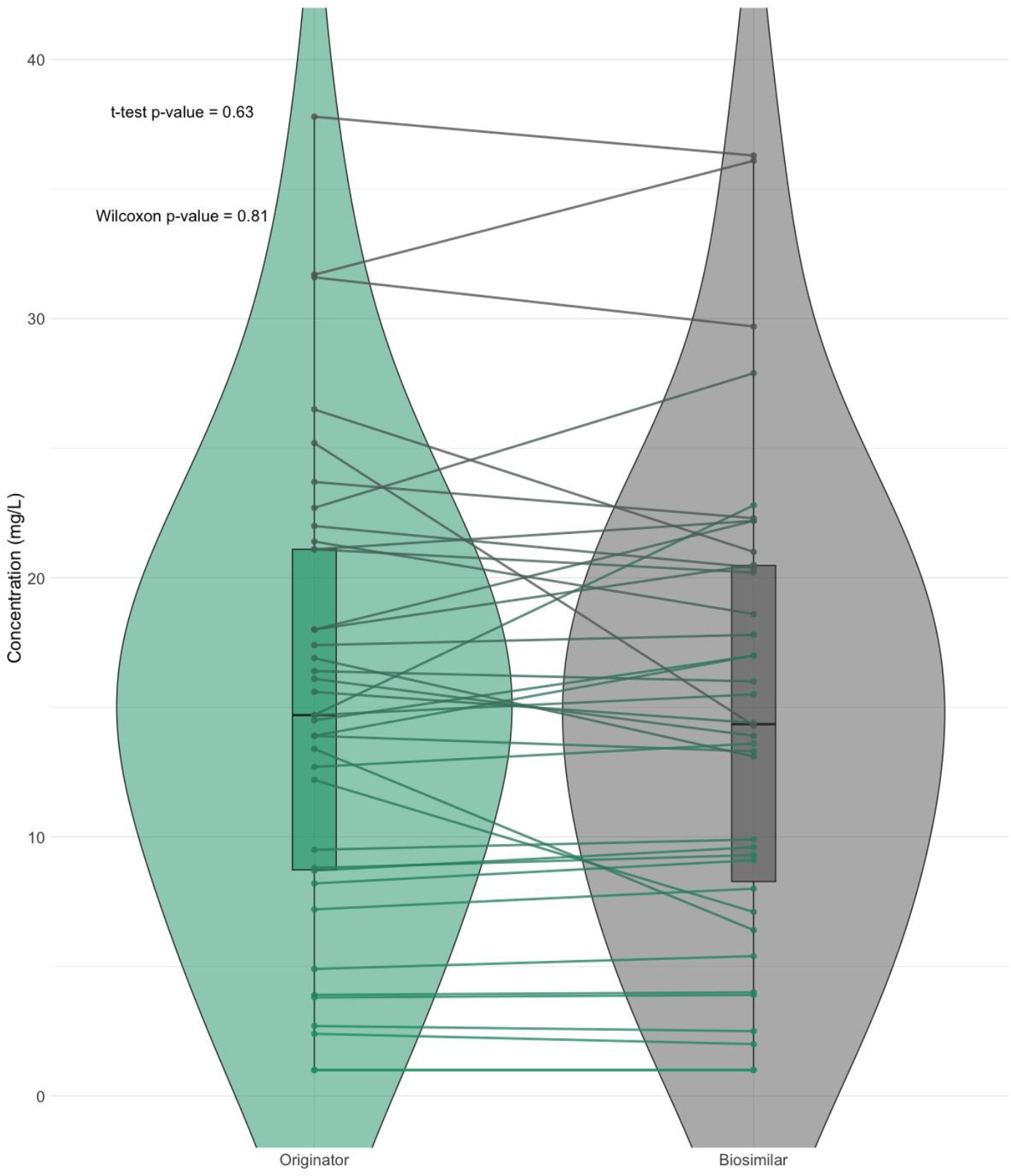
Serum levels of natalizumab after the last infusion of the originator and after the first infusion of the biosimilar.

**Figure 2:**
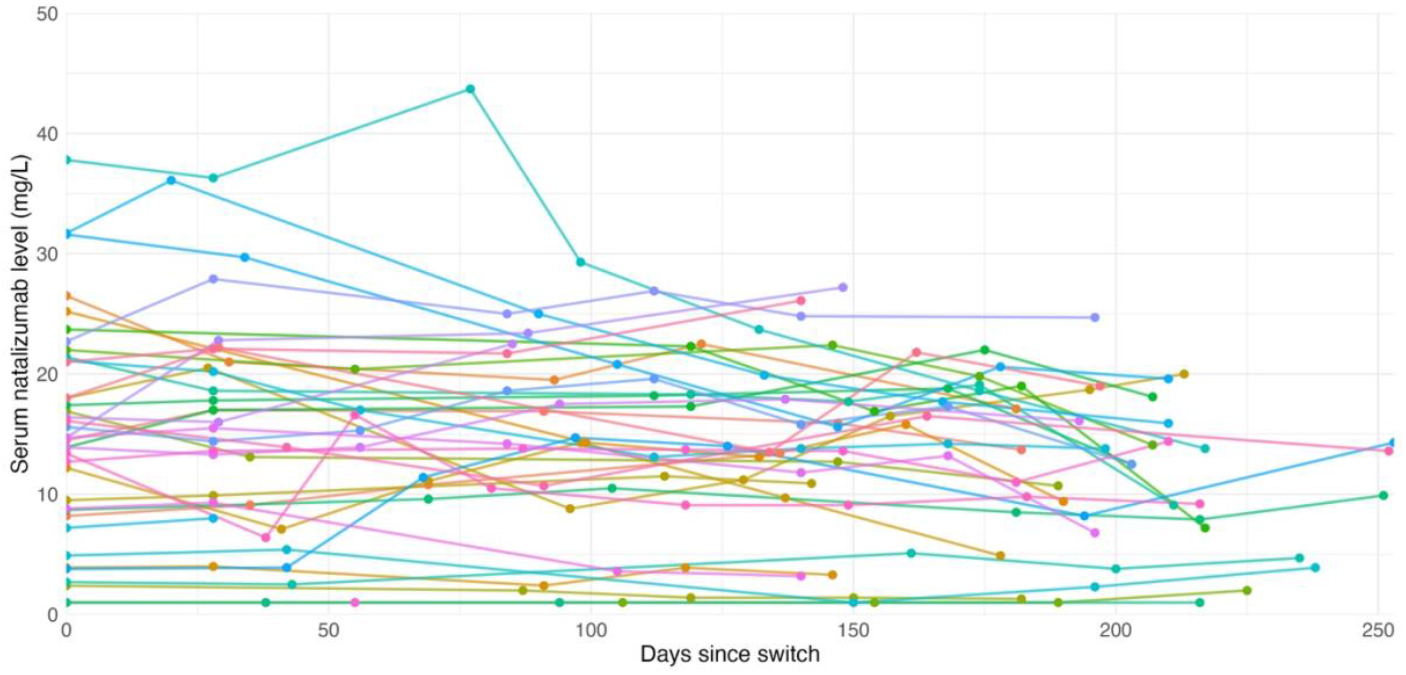
Longitudinal data for all persons with MS with repeated serum levels of natalizumab, with individual lines for each participant.

### Anti-drug antibodies

In this study ADAb were measured in the pwMS with low s-NTZ (less than 5 mg/L), with an in-house bridging assay automated on the AutoDELFIA platform. Biotinylated F(ab’)_2_ fragments of NTZ were used as solid phase reagent, and europium-labelled whole NTZ was used as tracer reagent. Results were considered positive for ADAb if signals were ten times higher than the assay blank.

### Anti-JCV antibody levels

Until December 31^st^, 2023, the anti-JCV antibody levels were analysed using the Stratify platform at Unilabs in Copenhagen at the expense of Biogen. No anti-JCV antibody test was available between January 1^st^, and March 31^st^, 2024. From March 31^st^, 2024, the samples were analysed using the Immunowell at the Medicover laboratory in Romania. In accordance with the reports from the assay platforms, we regarded antibody levels ≥0.4 in Stratify and ≥0.5 on the Immunowell platform as positive. For samples with an intermediate index value (0.2-0.4 for Stratify and 0.3-0.5 for Immunowell) inhibition tests were performed by the laboratories to determine whether the result was positive or negative (https://stratifyjcv.unilabsweb.com). For both tests a positive anti-JCV antibody index ≤0.8 was considered low, and an index >0.8 was considered high.

### Statistical analyses

All statistical analyses and data visualisations were performed using R (version 4.4.2). The main packages utilised for this analysis included *ggplot2* for data visualisation and *stats* for conducting the Shapiro-Wilk test and Student’s t-test. Statistical analyses were performed using the Student’s t-test to compare serum NTZ levels between groups. The normality of serum NTZ levels was assessed using the Shapiro-Wilk test. Differences in the proportion of JCV-positive cases across test platforms were assessed using a chi-square test. A p-value of less than 0.05 was considered statistically significant.

## Results

PwMS characteristics are presented in Table 1. Mean age was 44.5 years, 72% were female, mean disease duration was 14.9 years and median EDSS 2.0. Mean NTZ treatment duration was 10.4 years (range 3.7-17.0) before switching. Twelve pwMS were treatment naïve, while 23 had switched from low-efficacy DMTs, and four had switched from HE-DMT to NTZ. We found no significant changes in leukocyte levels before and after switching to biosimilar NTZ, as shown in Table 2.

**Table 1:**
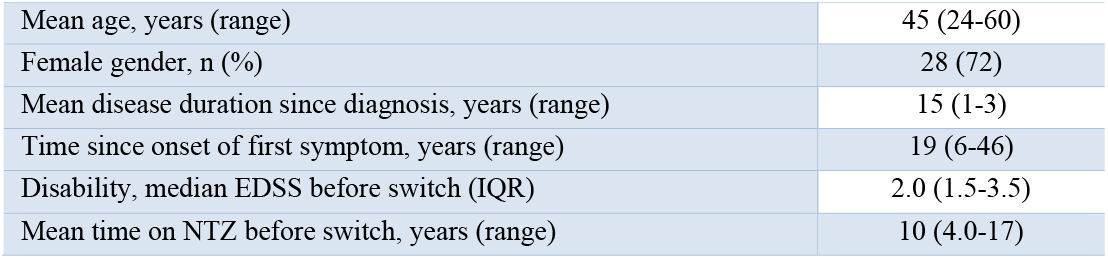
Demographic and clinical characteristics.

**Table 2:**
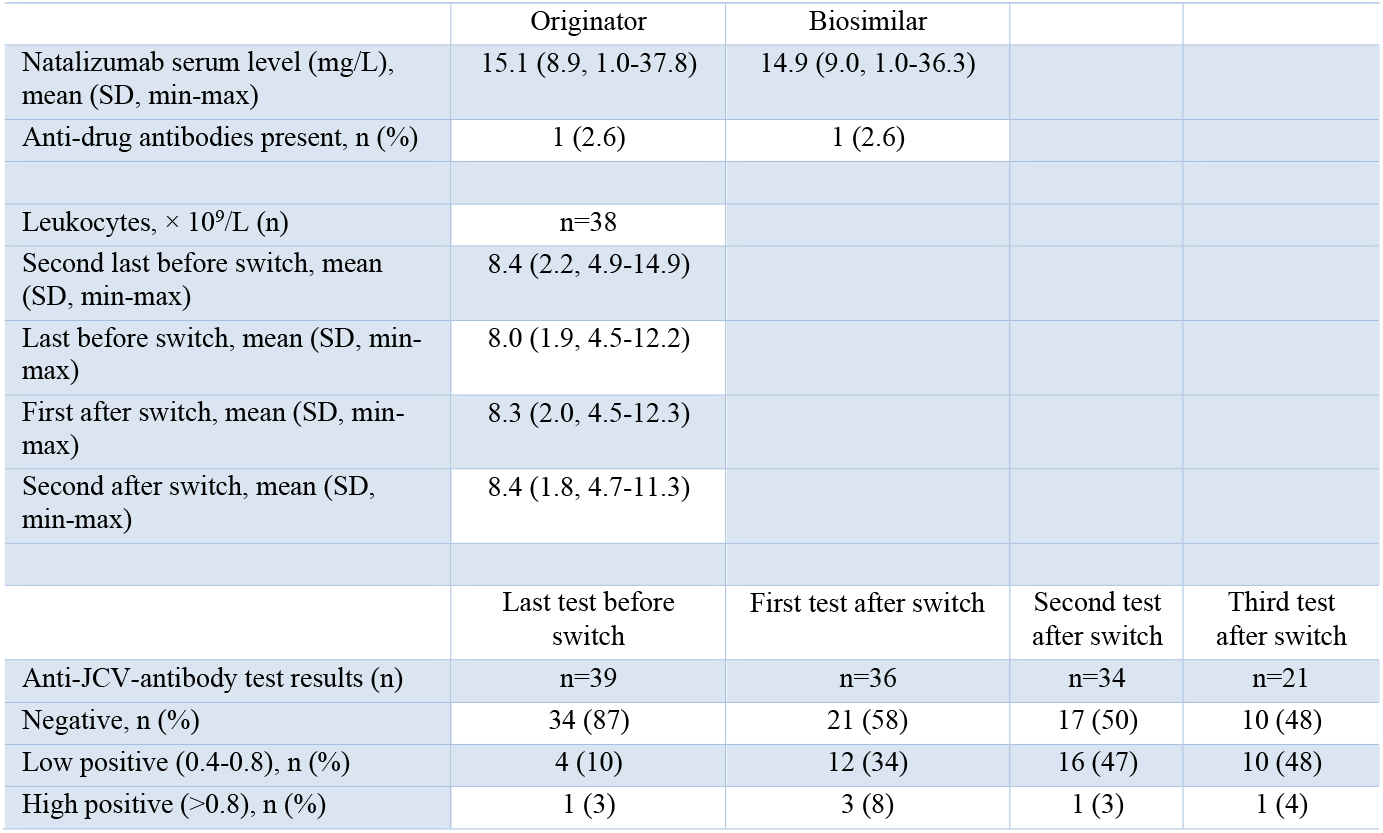
Natalizumab serum levels, ADAb, leukocytes and JCV-antibody index.

S-NTZ were 15.1 mg/L (SD 8.9, range 1.0-37.8) before switching and 14.9 mg/L (SD 9.0, range 1.0-36.3) after switching (mean difference 0.2 mg/L, p=0.63) (Figure 1). In the observed period infusion intervals were adjusted in some pwMS based on s-NTZ level, in others due to side-effects, and these were repeatedly tested.

Eleven pwMS reported new side effects during the first eight months after switch to biosimilar, whereof nine reported side effects after the first infusion, either the day after infusion or gradually during the first month after switch, and two pwMS reported side effects after the third infusion. The most common reported side effects were increased levels of fatigue, headache and pain, as well as lack of what some pwMS describe as a boost experienced after infusion with the originator. Five of the pwMS reporting side effects had s-NTZ levels of more than 25.0 mg/L. They were treated with extended interval dosing, leading to a reduction, but not complete attenuation of the symptoms. None of the pwMS changed DMT because of the side effects and no new symptomatic treatment was initiated due to side effects during the first eight months. One pwMS switched DMT after nine months because of side effects and one after 12 months due to both possible side effect and increasing JCV antibody level. One pwMS with a stable disease on NTZ for 12 years experienced a relapse a few days after receiving the second infusion with biosimilar NTZ, coincidently also four months after extending dose interval to six weeks.

When dividing the cohort in one group with reported side effects (n=11) and one without (n=28), the analysis of s-NTZ levels before the first infusion with biosimilar revealed a significant difference between the two groups (t = 2.23, p = 0.04). The mean s-NTZ level before switch for the group experiencing side effects from switching was 20.7 mg/L (range 1.0– 37.8 mg/L, SD 11.0), while the group without side effects had a mean level of 12.9 mg/L (range: 1.0–22.8 mg/L, SD 7.6). Before the second infusion the group with side effects had a mean level of 19.7 mg/L (range: 1.0–36.1 mg/L, SD = 10.4), while the group without side effects had a mean level of 12.5 mg/L (range: 1.0–27.9 mg/L, SD 8.4), where the difference between the groups was not statistically significant (t = 2.01, p = 0.06) .

An overview of the anti-JCV antibody index is shown in Figure 3, and the proportion of pwMS with low and high index in the tests is shown in Table 2. Of the pwMS 13% had a positive anti-JCV antibody index prior to the switch (Stratify test), while 42%, 50% and 52% of the pwMS had positive test results on average 4.6 (range 3.5-6.9), 6.4 (range 4.8-10.4) and 8.6 (6.0-11.1) months after switching (Immunowell test). The proportion of positive anti-JCV antibody index was significantly higher after switching from the Stratify platform to the Immunowell platform (p = 2.1 x 10^-3^), Four pwMS stopped NTZ treatment because of high anti-JCV antibody index after switching. One pwMS was lost to follow up after two infusions of biosimilar NTZ.

**Figure 3:**
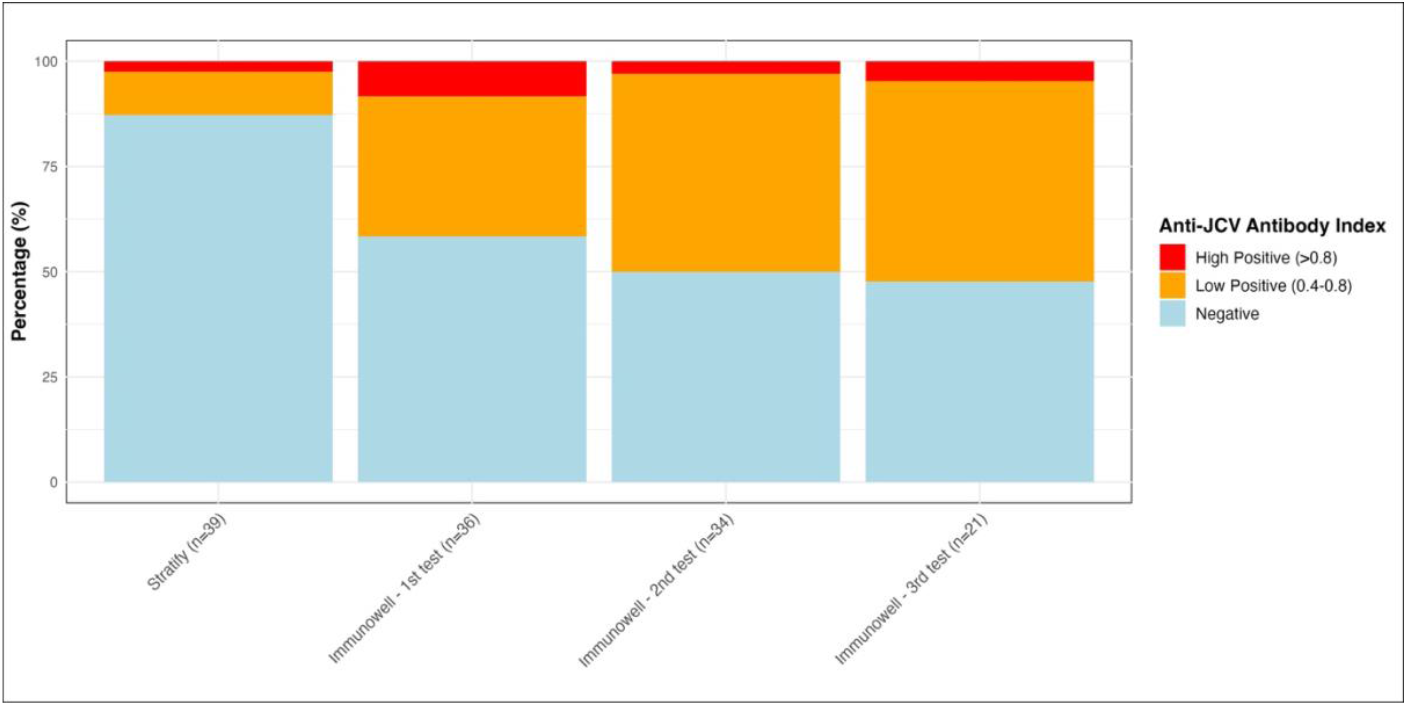
Anti-JCV antibody results with the originator (Stratify) and biosimilar (Immunowell) companion diagnostic tests.

## Discussion

Both s-NTZ levels and the proportion with ADAb were stable before and after switching from originator to biosimilar NTZ. However, we identified challenges with an increase in self-reported side effects, and an increased proportion of anti-JCV antibody positive pwMS with the new test.

In this cohort, 28% reported new side effects during the first eight months after switching from originator to biosimilar NTZ. While these side effects were generally modest and did not lead to new symptomatic treatment or switch of DMT, the perceived side-effects of the biosimilar drug were different. This was surprising, as there were no differences in pwMS-reported side effects between originator and biosimilar NTZ in the randomised controlled trial leading to approval of biosimilar NTZ (9). Some of the side effects were attenuated with extended dosing interval, indicating a possible dose-response relationship. However, also pwMS with s-NTZ levels less than 25.0 mg/L reported side effects. There is a possibility that this may be a nocebo effect, caused by pwMS distress when becoming aware of the switch from originator to biosimilar.

Differences in immunogenicity has been a concern when switching patients from originator to biosimilar treatments. A difference in immunogenicity occurred when changing from originator to biosimilar erythropoietin, leading to increased formation of ADAb and reduced efficacy of the drug (12, 13). In MS, one study has indicated a difference in efficacy between originator and generic fingolimod, probably caused by differences in drug levels (14). However, in our population, the drug levels with originator and biosimilar NTZ were similar, and also only one patient had detectable ADAb. This is in line with previous findings for biosimilar monoclonal antibody drugs approved for other conditions, with safety, efficacy and side effects comparable to their reference medications (15).

With the switch from originator to biosimilar NTZ, the Stratify platform for anti-JCV antibody testing was no longer available. Historically, the risk of PML posed major challenges to the use of NTZ, and the anti-JCV antibody test by Stratify was sponsored by Biogen, as a companion diagnostic test, developed in parallel to the drug to ensure safety. The availability of this test was restricted to those treated with originator NTZ and therefore the biosimilar manufacturer provided a new companion diagnostic test on the Immunowell platform. We identified a large increase in the proportion of pwMS with positive anti-JCV antibody index over the course of the observation period, both in the low and high ranges coinciding with this change of test platform. We cannot completely rule out the possibility that this increase was caused by a local change in JCV exposure. However, it is much more likely that the change was caused by different properties between the test platforms. For clinicians, this change in JCV status has created challenges in calculating the risk of PML based on a test that has not been compared with the previous. Recently, two previously anti-JCV antibody negative pwMS who turned positive with the new platform and then negative when retested with the old platform were reported (16). We strongly support harmonisation of anti-JCV antibody testing across laboratories as a result of our study.

Clinicians using biosimilar NTZ face challenges when confirmatory testing for positive anti-JCV antibody results is unavailable for previously negative pwMS. In such cases, they must decide whether to discontinue NTZ, extend dosing intervals, or continue treatment despite the high risk in JCV-positive pwMS. Even though several reports have demonstrated successful treatment switches from NTZ to anti-CD20 treatments, switching treatment is associated with risks of rebound disease activity and side effects (17). As an alternative to treatment switch, extended interval dosing has been suggested to reduce the long-term risk of PML in pwMS with low anti-JCV antibody levels (6, 7, 18). One recent study has shown that extended interval dosing guided by therapeutic drug monitoring, may allow dosing intervals up to ten weeks with no safety signals (16), but this needs confirmation in large trials. Another recent study found up to a 30-fold difference in s-NTZ levels, consistent with our findings, and highlighting the potential need and benefit to personalise NTZ treatment (19).

Evaluating s-NTZ levels has proven valuable in our cohort, as levels above 25.0 mg/L seemed to be associated with an increased risk of side effects, which improved to some degree with extended dosing intervals. This approach allows clinicians and pwMS to personalise NTZ infusion intervals. Furthermore, one pwMS experienced a relapse a few days after receiving the second infusion with biosimilar NTZ, where low s-NTZ levels were detected. A relationship between low drug levels and lack of clinical efficacy has been demonstrated for other biologic drugs used for other indications but remains to be proven for NTZ in pwMS (20-22). We suggest that a threshold for minimum s-NTZ levels should be considered, with potential adjustments of treatment intervals. Thus both pwMS in the high and low range of s-NTZ levels may profit from therapeutic drug monitoring similar to other immune mediated inflammatory diseases (23-25).

It is important to note that the relationship between s-NTZ levels and side effects is complex, and other factors may play a role. In addition, an increased proportion of pwMS tested positive to anti-JCV antibodies with the biosimilar companion diagnostic test. The restrictions and lacking harmonisation of the current companion diagnostic tests for NTZ complicate the switch and in effect reduce access to biosimilar NTZ. While the investigations of s-NTZ levels and side effects warrant further studies in larger cohorts, the challenges related to lack of harmonisation of anti-JCV antibody tests is largely a regulatory problem that requires transparency and collaboration.

In conclusion, the switch from originator to biosimilar natalizumab appears safe in terms of drug levels and immunogenicity, but challenges remain with increased self-reported side effects and a higher proportion of positive anti-JCV antibody results with the new companion diagnostic test. These findings highlight the need for further studies on the clinical implications of s-NTZ levels and for harmonisation of anti-JCV antibody testing to support clinical decision-making and ensure safe and effective use of biosimilars.

## Acknowledgement

We thank all pwMS for their contribution to this study.

## Data Availability Statement

Due to privacy restrictions data are only available on request.

## References

1. Polman CH, O’Connor PW, Havrdova E, Hutchinson M, Kappos L, Miller DH, et al. A randomized, placebo-controlled trial of natalizumab for relapsing multiple sclerosis. 2006 2006.

2. He A, Merkel B, Brown JWL, Zhovits Ryerson L, Kister I, Malpas CB, et al. Timing of high-efficacy therapy for multiple sclerosis: a retrospective observational cohort study. Lancet Neurol. 2020;19(4):307–16.

3. Kobelt G, Thompson A, Berg J, Gannedahl M, Eriksson J. New insights into the burden and costs of multiple sclerosis in Europe. Multiple sclerosis (Houndmills, Basingstoke, England). 2017;23(8):1123–36.

4. Ho PR, Koendgen H, Campbell N, Haddock B, Richman S, Chang I. Risk of natalizumab-associated progressive multifocal leukoencephalopathy in patients with multiple sclerosis: a retrospective analysis of data from four clinical studies. Lancet Neurol. 2017;16(11):925–33.

5. Oshima Y, Tanimoto T, Yuji K, Tojo A. Drug-associated progressive multifocal leukoencephalopathy in multiple sclerosis patients. Mult Scler. 2018:1352458518786075.

6. Foley JF, Defer G, Ryerson LZ, Cohen JA, Arnold DL, Butzkueven H, et al. Comparison of switching to 6-week dosing of natalizumab versus continuing with 4-week dosing in patients with relapsing-remitting multiple sclerosis (NOVA): a randomised, controlled, open-label, phase 3b trial. Lancet Neurol. 2022;21(7):608–19.

7. Perncezky J, Sellner J. Natalizumab extended-interval dosing in multiple sclerosis to mitigate progressive multifocal leukoencephalopathy risk: initial study evidence and real-world experience. J Cent Nerv Syst Dis. 2022;14:11795735221135485.

8. Brun MK, Gehin JE, Bjørlykke KH, Warren DJ, Klaasen RA, Sexton J, et al. Clinical consequences of infliximab immunogenicity and the effect of proactive therapeutic drug monitoring: exploratory analyses of the randomised, controlled NOR-DRUM trials. Lancet Rheumatol. 2024;6(4):e226–e36.

9. Hemmer B, Wiendl H, Roth K, Wessels H, Höfler J, Hornuss C, et al. Efficacy and Safety of Proposed Biosimilar Natalizumab (PB006) in Patients With Relapsing-Remitting Multiple Sclerosis: The Antelope Phase 3 Randomized Clinical Trial. JAMA Neurol. 2023;80(3):298–307.

10. Kurtzke JF. Rating neurologic impairment in multiple sclerosis: an expanded disability status scale (EDSS). Neurology. 1983;33(11):1444–52.

11. Sorensen PS, Jensen PE, Haghikia A, Lundkvist M, Vedeler C, Sellebjerg F, et al. Occurrence of antibodies against natalizumab in relapsing multiple sclerosis patients treated with natalizumab. Multiple sclerosis (Houndmills, Basingstoke, England). 2011;17(9):1074–8.

12. Kuhlmann M, Marre M. Lessons learned from biosimilar epoetins and insulins. The British Journal of Diabetes & Vascular Disease. 2010;10(2):90–7.

13. Praditpornsilpa K, Tiranathanagul K, Kupatawintu P, Jootar S, Intragumtornchai T, Tungsanga K, et al. Biosimilar recombinant human erythropoietin induces the production of neutralizing antibodies. Kidney Int. 2011;80(1):88–92.

14. Okuda DT, Tardo LM, Wright CM, Munoz SB, Punnen TG, Patel MA, et al. Clinical and radiological implications of subpotent generic fingolimod in multiple sclerosis: a case series. Ther Adv Neurol Disord. 2024;17:17562864241300047.

15. Jorgensen KK, Olsen IC, Goll GL, Lorentzen M, Bolstad N, Haavardsholm EA, et al. Switching from originator infliximab to biosimilar CT-P13 compared with maintained treatment with originator infliximab (NOR-SWITCH): a 52-week, randomised, double-blind, non-inferiority trial. Lancet. 2017;389(10086):2304–16.

16. Toorop AA, van Lierop ZY, Gelissen LM, Hoitsma E, Zeinstra EM, van Rooij LC, et al. Prospective trial of natalizumab personalised extended interval dosing by therapeutic drug monitoring in relapsing-remitting multiple sclerosis (NEXT-MS). Journal of Neurology, Neurosurgery & Psychiatry. 2024;95(5):392–400.

17. Brown JD, Muston BT, Massey J. Switching from natalizumab to an anti-CD20 monoclonal antibody in relapsing remitting multiple sclerosis: A systematic review. Multiple Sclerosis and Related Disorders. 2024;86.

18. Ryerson LZ, Foley J, Chang I, Kister I, Cutter G, Metzger RR, et al. Risk of natalizumab-associated PML in patients with MS is reduced with extended interval dosing. Neurology. 2019;93(15):e1452–e62.

19. Moskorova D, Kacirova I, Hradilek P, Matlak P, Brozmanova H, Kusnierova P, et al. Title: Analysis of serum natalizumab concentrations obtained during routine clinical care in patients with multiple sclerosis: a cross-sectional study. Multiple sclerosis and related disorders.

20. Krieckaert CL, van Tubergen A, Gehin JE, Hernandez-Breijo B, Le Meledo G, Balsa A, et al. EULAR points to consider for therapeutic drug monitoring of biopharmaceuticals in inflammatory rheumatic and musculoskeletal diseases. Ann Rheum Dis. 2023;82(1):65–73.

21. Adedokun OJ, Sandborn WJ, Feagan BG, Rutgeerts P, Xu Z, Marano CW, et al. Association between serum concentration of infliximab and efficacy in adult patients with ulcerative colitis. Gastroenterology. 2014;147(6):1296-307.e5.

22. Vande Casteele N, Khanna R, Levesque BG, Stitt L, Zou GY, Singh S, et al. The relationship between infliximab concentrations, antibodies to infliximab and disease activity in Crohn’s disease. Gut. 2015;64(10):1539–45.

23. Syversen SW, Goll GL, Jorgensen KK, Olsen IC, Sandanger O, Gehin JE, et al. Therapeutic drug monitoring of infliximab compared to standard clinical treatment with infliximab: study protocol for a randomised, controlled, open, parallel-group, phase IV study (the NOR-DRUM study). Trials. 2020;21(1):13.

24. Syversen SW, Goll GL, Jorgensen KK, Sandanger O, Sexton J, Olsen IC, et al. Effect of Therapeutic Drug Monitoring vs Standard Therapy During Infliximab Induction on Disease Remission in Patients With Chronic Immune-Mediated Inflammatory Diseases: A Randomized Clinical Trial. JAMA. 2021;325(17):1744–54.

25. Syversen SW, Jørgensen KK, Goll GL, Brun MK, Sandanger Ø, Bjørlykke KH, et al. Effect of Therapeutic Drug Monitoring vs Standard Therapy During Maintenance Infliximab Therapy on Disease Control in Patients With Immune-Mediated Inflammatory Diseases: A Randomized Clinical Trial. JAMA. 2021;326(23):2375–84.

